# Bibliometric Analysis of Academic Journal Articles Reporting Results of Psychedelic Clinical Studies

**DOI:** 10.1101/2021.11.22.21266718

**Authors:** Jeremy Weleff, Teddy J. Akiki, Brian S. Barnett

**Author notes:** Corresponding author: Jeremy Weleff, DO.

## Abstract

After a decades long period of investigational dormancy, there is renewed interest in employing psychedelics as treatments for mental illness and addiction. The academic journals, journal articles, academic institutions, and countries that have helped sustain clinical psychedelic research and the evolution of the literature on clinical studies of psychedelic compounds have only been minimally investigated. Therefore, in we conducted a bibliometric analysis of clinical studies of 5-methoxy-N, N-dimethyltryptamine (5-MeO-DMT), ayahuasca, dimethyltryptamine (DMT), lysergic acid diethylamide (LSD), ibogaine, mescaline, 3,4-methylenedioxymethamphetamine (MDMA), and psilocybin published from 1965-2018. Our search revealed 320 articles published across 106 journals. After a nearly quarter century lull between the 1970s and 1990s, publications in this area have resurged over the last two decades and continue on an upward trajectory, with most clinical studies now focusing on LSD, MDMA, and psilocybin. A subanalysis of the ten most cited articles in psychedelic research prior to 2010 and afterwards demonstrated a shift from research on risks of psychedelics, primarily those of MDMA, to research on therapeutic applications, predominantly those of psilocybin. We also conducted network analyses of inter-country collaborations in psychedelic research, which suggested that psychedelic researchers in the United Kingdom have more diverse international collaborations.

## INTRODUCTION

After a decades long period of clinical and investigational dormancy, there is renewed interest in employing psychedelics as treatments for mental illness and addiction (Murnane 2018; Sessa 2018; Kelly et al. 2019). At the forefront of investigations into the therapeutic applications of psychedelics are 3,4-methylenedioxymethamphetamine (MDMA), currently in phase 3 trials for treatment of posttraumatic stress disorder, and psilocybin, amid phase 2 trials for treatment of both major depressive disorder and treatment resistant depression. Given the treatment potential demonstrated by therapeutic approaches involving these compounds, the United States (US) Food and Drug Administration has bestowed a Breakthrough Therapy designation on psilocybin-assisted psychotherapy (Compass Pathways 2018; Business Wire 2019) and MDMA-assisted psychotherapy (Burge 2017). While regulatory, financial, and cultural issues remain important barriers to further advances in psychedelic medicine; researchers, psychedelic-focused non-profit organizations, philanthropists, and others continue to make significant strides in bringing these compounds back from the brink of scientific dismissal and into the clinic once more.

The evolution of the literature on clinical studies of psychedelics has only been studied minimally (Lawrence et al. 2021), meaning we know little about the people, articles, and institutions that have helped sustain and continue to propel clinical psychedelic research forward. In order to further our understanding of the clinical psychedelic literature, we performed a bibliometric analysis of journal articles reporting findings from clinical studies of the prominent psychedelics, lysergic acid diethylamide (LSD), psilocybin, dimethyltryptamine (DMT), 5-methoxy-N, N-dimethyltryptamine (5-MeO-DMT), ibogaine, mescaline, and MDMA (ecstasy) published from 1965-2018.

## METHODS

### Search strategy

On November 24^th^, 2020, we searched PubMed using the “All Fields” search capability for journal articles published from1936-2020 containing the following search terms: (*“psilocybin”) OR (“LSD”) OR (“lysergic acid diethylamide”) OR (“mescaline”) OR (“MDMA”) OR (“ecstasy”) OR (“DMT”) OR (“dimethyltryptamine”) OR (“5-methoxy-N, N-dimethyltryptamine”) OR (“5-MeO-DMT”) OR (“ibogaine”)*. We limited results to clinical studies involving humans using PubMed’s built-in filtering functionality. We then entered the PubMed IDs (PMIDs) of resulting articles into the Clarivate *Web of Science* (WoS) Core Collection for all available years (1965-2020) and extracted detailed metadata on the articles. Though we had initially planned to evaluate relevant clinical studies published through 2020, given the sometimes substantial delays in indexing articles in PubMed (Irwin and Rackham 2017), we later decided to exclude papers published after 2018 to maximize the integrity of the dataset.

After exporting results from WoS, authors JW and BB next evaluated article abstracts and, when necessary, the articles themselves to determine whether the articles reported findings of clinical studies investigating the effects of the psychedelics in humans and were published in English. Discrepancies about whether to exclude an article were resolved through discussion between the authors. Notably, the authors found that a number of search terms overlapped with other commonly used acronyms in medical research (for example, the search term “DMT” resulted in articles on “disease modifying therapy” in the treatment of multiple sclerosis and “LSD” produced results for papers mentioning the statistical terms “least square difference” and “least significant difference”), leading to numerous articles being excluded from the final dataset.

### Bibliometric analysis

Bibliometric analysis allows for quantification of publication trends and provides the tools necessary for dissecting and categorizing the academic papers fueling those trends. We used *bibliometrix* (Aria and Cuccurullo 2017), an open-source R-tool designed to perform comprehensive science mapping analyses, and its associated web-based app *biblioshiny* to analyze articles detailing clinical studies of psychedelics and extract information about the journals they were published in, their year of publication, their citation count, and their authors (including authors’ academic institutions and the countries of those institutions).

We also conducted a network analysis to study the pattern of co-author collaboration across countries (Aria and Cuccurullo 2017). A co-author collaboration network based on affiliation country was generated by indexing the co-occurrence of countries in the author list of an article. To quantitatively describe a country’s propensity to engage in collaborations, we calculated a nodal metric of *betweenness centrality* that measures how often a node appears on the shortest path between nodes (Brandes 2001). Centrality values were then min-max normalized to the range [0, 1] for simplicity. We used Gephi 0.9.2 (https://github.com/gephi/gephi) and a force-directed layout algorithm for network visualization (Bastian, Heymann, and Jacomy 2009; Jacomy et al. 2014).

## RESULTS

696 articles were identified during our initial PubMed search. 62 articles were excluded due to not being indexed in WoS and 37 for being published after 2018. An additional 277 were excluded for various other reasons, detailed in Figure 1, resulting in a final dataset of 320 articles published across 106 journals and authored by 900 individuals. The mean number of years since article publication was 17.2 ± 14.5 (mean ± standard deviation) years prior to search and mean citation count per article was 61.5 ± 72.2. Articles reporting findings of clinical studies published during the first wave of psychedelic research reached a peak of six articles annually in (1966 and 1968), while only four articles in total were published from 1977-1993. Recent publication remains on an upward trend and reached an annual peak of 23 papers published during 2018. Figure 2 illustrates annual publication of articles from 1965-2018 and trends in publications on specific substances over this period. Articles most frequently reported findings from clinical studies of MDMA (172), followed by psilocybin (57), LSD (54), DMT (17), ayahuasca (19), ibogaine (10), and mescaline (6). Using the definition of the “Recent Cohort” (published after 2010) of psychedelic studies (Lawrence et al. 2021), 93 MDMA (54.1% of studies on this compound), 32 LSD (59.3% of studies on this compound), 21 psilocybin (36.8% of studies on this compound), 15 DMT (88.2% of studies on this compound), 11 ayahuasca (57.9% of studies on this compound), 6 ibogaine (60% of studies on this compound), and 6 mescaline (100% of studies on this compound) studies were found to be published from 2010 to 2018. The ten most cited articles from both the recent and older cohorts of psychedelic studies are listed in Table 1.

**Table 1.** Top 10 most cited psychedelic clinical study articles by old and new cohort of psychedelic research

| Top 10 most cited studies from the old cohort of psychedelic research (before 2010) |  |  |  |  |  |
| --- | --- | --- | --- | --- | --- |
|  | Title | Authors | Journal | Year | Citations |
| 1 | Positron emission tomographic evidence of toxic effect of MDMA (ecstasy) on brain serotonin neurons in human beings | McCann, UD; Szabo, Z; Scheffel, U; Dannals, RF; Ricaurte, GA | *Lancet* | 1998 | 486 |
| 2 | Psilocybin can occasion mystical-type experiences having substantial and sustained personal meaning and spiritual significance | Griffiths, R. R.; Richards, W. A.; McCann, U.; Jesse, R. | *Psychopharmacology* | 2006 | 424 |
| 3 | Psilocybin induces schizophrenia-like psychosis in humans via a serotonin-2 agonist action | Vollenweider, FX; Vollenweider-Scherpenhuyzen, MFI; Babler, A; Vogel, H; Hell, D | *Neuroreport* | 1998 | 406 |
| 4 | Psychological and cardiovascular effects and short-term sequelae of mdma (ecstasy) in MDMA-naive healthy volunteers | Vollenweider, FX; Gamma, AG; Liechti, M; Huber, T | *Neuropsychopharmacology* | 1998 | 277 |
| 5 | Serotonin neurotoxicity after (+/-)3,4-methylenedioxymethamphetamine (MDMA; "ecstasy") - a controlled-study in humans | Mccann, Ud; Ridenour, A; Shaham, Y; Ricaurte, Ga | *Neuropsychopharmacology* | 1994 | 259 |
| 6 | Non-linear pharmacokinetics of MDMA ('ecstasy') in humans | de la Torre, R; Farre, M; Ortuno, J; Mas, M; Brenneisen, R; Roset, PN; Segura, J; Cami, J | *British Journal Of Clinical Pharmacology* | 2000 | 244 |
| 7 | Recreational use of ecstasy (MDMA) is associated with elevated impulsivity | Morgan, MJ | *Neuropsychopharmacology* | 1998 | 232 |
| 8 | Cardiovascular and neuroendocrine effects and pharmacokinetics of 3,4-methylenedioxymethamphetamine in humans | Mas, M; Farre, M; De la Torre, R; Roset, PN; Ortuno, J; Segura, J; Cami, J | *Journal Of Pharmacology And Experimental Therapeutics* | 1999 | 231 |
| 9 | Dose-response study of N,N-dimethyltryptamine in humans .2. subjective effects and preliminary-results of a new rating-scale | Strassman, Rj; Qualls, Cr; Uhlenhuth, Eh; Kellner, R | *Archives Of General Psychiatry* | 1994 | 219 |
| 10 | Gender differences in the subjective effects of MDMA | Liechti, ME; Gamma, A; Vollenweider, FX | *Psychopharmacology* | 2001 | 194 |
| Top 10 most cited studies from the new cohort of psychedelic research (after 2010) |  |  |  |  |  |
| 1 | Pilot study of psilocybin treatment for anxiety in patients with advanced-stage cancer | Grob, Charles S.; Danforth, Alicia L.; Chopra, Gurpreet S.; Hagerty, | *Archives Of General Psychiatry* | 2011 | 345 |
|  |  | Marycie; Mckay, Charles R.; Halberstadt, Adam L.; Greer, George R. |  |  |  |
| 2 | Neural correlates of the psychedelic state as determined by fMRI studies with psilocybin | Carhart-Harris, Robin L.; Erritzoe, David; Williams, Tim; Stone, James M.; Reed, Laurence J.; Colasanti, Alessandro; Tyacke, Robin J.; Leech, Robert; Malizia, Andrea L.; Murphy, Kevin; Hobden, Peter; Evans, John; Feilding, Amanda; Wise, Richard G.; Nutt, David J. | *Proceedings Of The National Academy Of Sciences Of The United States Of America* | 2012 | 323 |
| 3 | Psilocybin produces substantial and sustained decreases in depression and anxiety in patients with life-threatening cancer: a randomized double-blind trial | Griffiths, Roland R.; Johnson, Matthew W.; Carducci, Michael A.; Umbricht, Annie; Richards, William A.; Richards, Brian D.; Cosimano, Mary P.; Klinedinst, Margaret A. | *Journal Of Psychopharmacology* | 2016 | 292 |
| 4 | Psilocybin with psychological support for treatment-resistant depression: an open-label feasibility study | Carhart-Harris, Robin L.; Bolstridge, Mark; Rucker, James; Day, Camilla M. J.; Erritzoe, David; Kaelen, Mendel; Bloomfield, Michael; Rickard, James A.; Forbes, Ben; Feilding, Amanda; Taylor, David; Pilling, Steve; Curran, Valerie H.; Nutt, David J. | *Lancet Psychiatry* | 2016 | 286 |
| 5 | Rapid and sustained symptom reduction following psilocybin treatment for anxiety and depression in patients with life-threatening cancer: a randomized controlled trial | Ross, Stephen; Bossis, Anthony; Guss, Jeffrey; Agin-Liebes, Gabrielle; Malone, Tara; Cohen, Barry; Mennenga, Sarah E.; | *Journal Of Psychopharmacology* | 2016 | 268 |
|  |  | Belser, Alexander;<br>Kalliontzi,<br>Krystallia; Babb,<br>James; Su, Zhe;<br>Corby, Patricia;<br>Schmidt, Brian L. |  |  |  |
| 6 | The safety and efficacy of +/- 3,4-methylenedioxymethamphetamine-assisted psychotherapy in subjects with chronic, treatment-resistant posttraumatic stress disorder: the first randomized controlled pilot study | Mithoefer, Michael C.; Wagner, Mark T.; Mithoefer, Ann T.; Jerome, Lisa; Doblin, Rick | *Journal Of Psychopharmacology* | 2011 | 262 |
| 7 | Psilocybin-assisted treatment for alcohol dependence: a proof-of-concept study | Bogenschutz, Michael P.; Forcehimes, Alyssa A.; Pommy, Jessica A.; Wilcox, Claire E.; Barbosa, P. C. R.; Strassman, Rick J. | *Journal Of Psychopharmacology* | 2015 | 233 |
| 8 | Psilocybin occasioned mystical-type experiences: immediate and persisting dose-related effects | Griffiths, Roland R.; Johnson, Matthew W.; Richards, William A.; Richards, Brian D.; Mccann, Una; Jesse, Robert | *Psychopharmacology* | 2011 | 227 |
| 9 | Pilot study of the 5-HT<sub>2A</sub>R agonist psilocybin in the treatment of tobacco addiction | Johnson, Matthew W.; Garcia-Romeu, Albert; Cosimano, Mary P.; Griffiths, Roland R. | *Journal Of Psychopharmacology* | 2014 | 203 |
| 10 | Safety and efficacy of lysergic acid diethylamide-assisted psychotherapy for anxiety associated with life-threatening diseases | Gasser, Peter; Holstein, Dominique; Michel, Yvonne; Doblin, Rick; Yazar-Klosinski, Berra; Passie, Torsten; Brenneisen, Rudolf | *Journal Of Nervous And Mental Disease* | 2014 | 194 |

**Figure 1:**
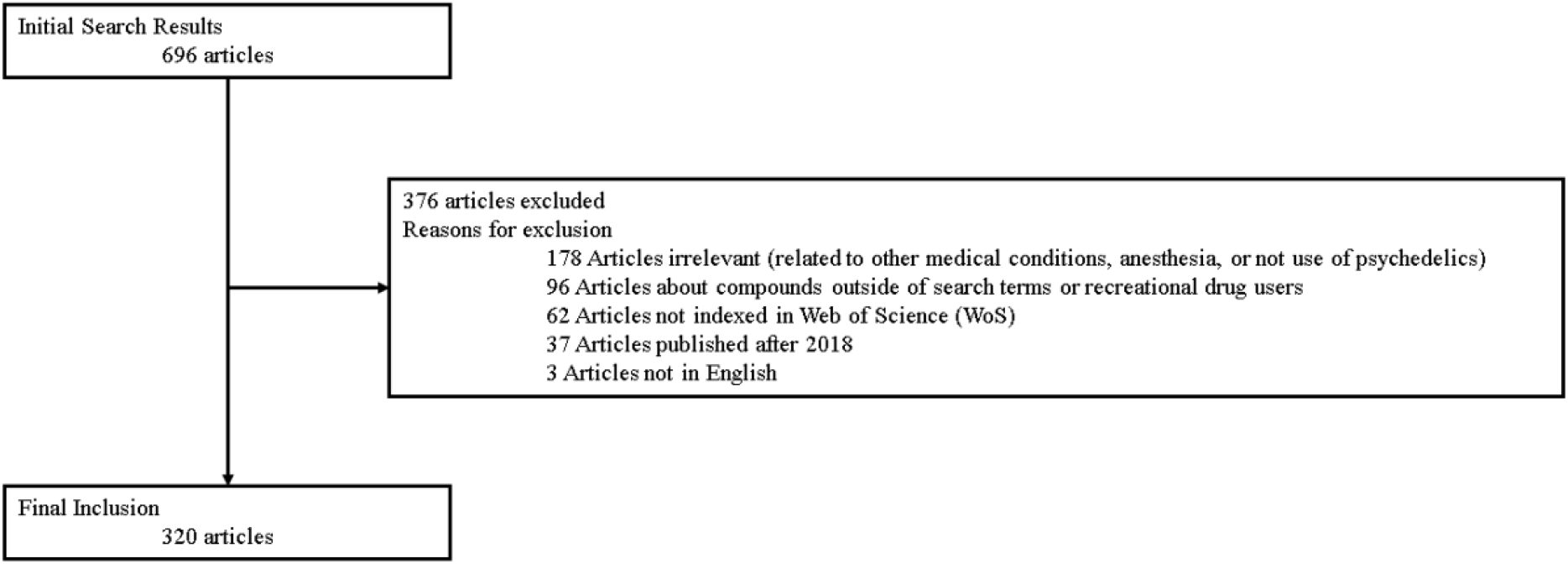
Article selection flowchart

**Figure 2.**
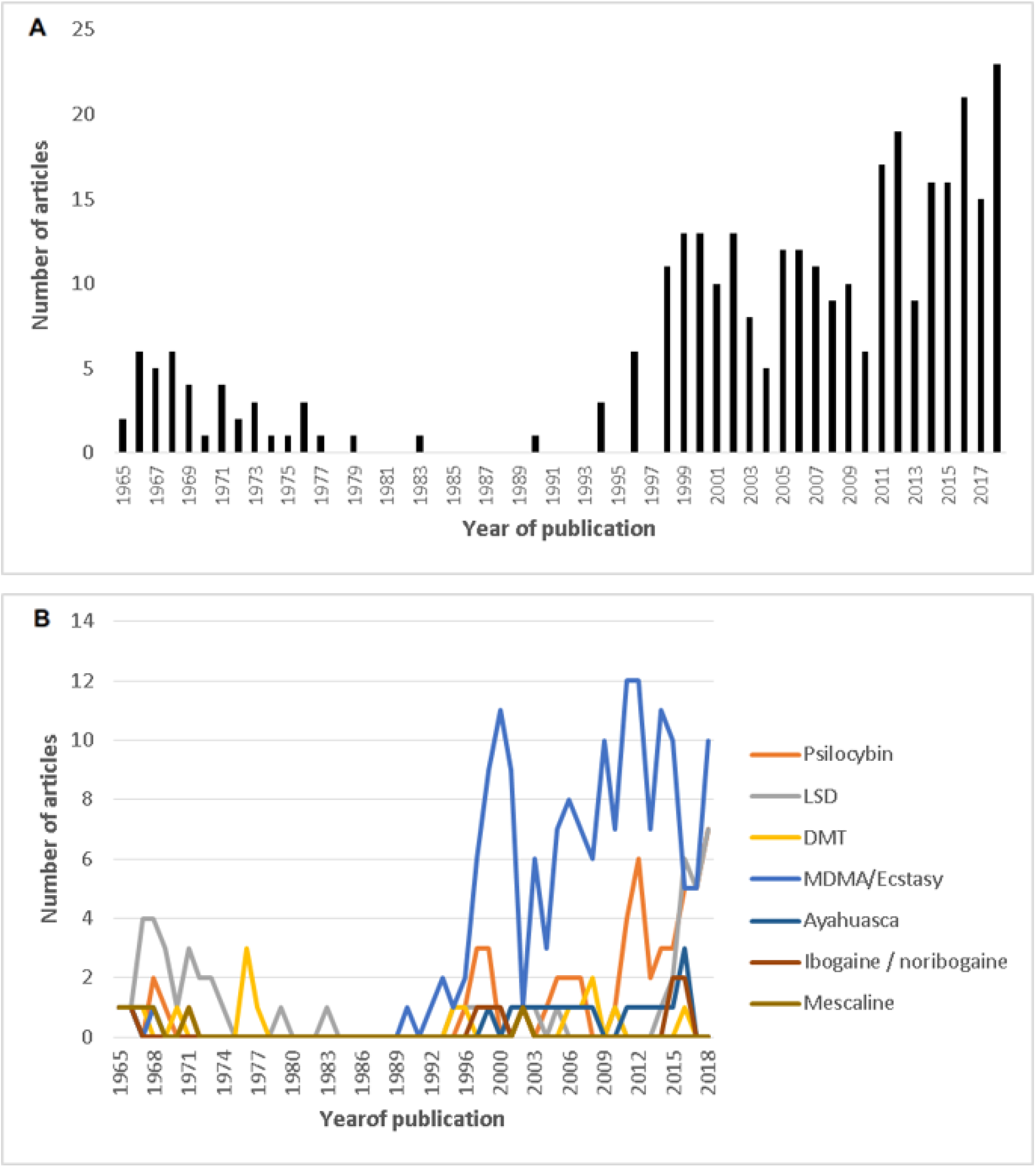
(A) Number of psychedelic clinical study articles published annually from 1965-2018. (B) Number of psychedelic clinical study articles published per year per compound.

### Journals

The journals publishing the most psychedelic clinical trial articles from 1965-2018 were: *Psychopharmacology* (55), *Journal of Psychopharmacology* (35), *Neuropsychopharmacology* (23) and *Biological Psychiatry* (10). Looking at publications during the “Recent Cohort”, the journals publishing most on psychedelics were *Psychopharmacology* (30; 54.5% of publications on psychedelics by this journal), *Journal of Psychopharmacology* (24; 68.6% of publications on psychedelics by this journal), *Neuropsychopharmacology* (9; 39.1% of publications on psychedelics by this journal) and *International Journal of Neuropsychopharmacology* (7; 100% of publications on psychedelics by this journal). The journals with the most citations of articles reporting findings from psychedelic clinical studies were *Psychopharmacology* (1,072), *Neuropsychopharmacology* (645), *Journal of Psychopharmacology* (595), and *Biological Psychiatry* (420). Further details on the number of relevant articles in journals publishing on psychedelics from 1965-2018, total citations for these journals, and the changes in number of publications on psychedelics in these journals over time are provided in the online supplement.

### Academic Institutions and Countries of Corresponding Authors

The academic institutions most frequently affiliated with corresponding authors were University of Basel (76), University of Zurich (58), Universitat Autònoma de Barcelona (36), Maastricht University (22), Johns Hopkins University (21), Pompeu Fabra University (19), and Imperial College London (16). Corresponding authors were most commonly based in Switzerland (70), US (64), Spain (38), United Kingdom (30) and the Netherlands (30). Countries (by all authors) with the greatest number of paper citations were the US (6,006), Switzerland (5,117), Spain (2,392), United Kingdom (2,366), Netherlands (1,078), Germany (1,073) and Australia (197). Among countries producing at least five clinical studies, the highest number of average article citations came from the US (93.8), United Kingdom (78.9), Switzerland (73.1), Spain (62.9), Germany (44.7), and Netherlands (35.9).

The country collaboration network initially consisted of 22 nodes. Three nodes (Czech Republic, Lithuania, and Israel) did not have connections and were removed, resulting in a 19-node connected network (Figure 3A). The node representing the United Kingdom has the largest number of connections to other nodes and occupies a central position in the network (Figure 3A-B). Other nodes, such as Switzerland, USA, and Germany have a stronger connection between them (Figure 3A), but a lower overall centrality in the network (Figure 3A-B).

**Figure 3.**
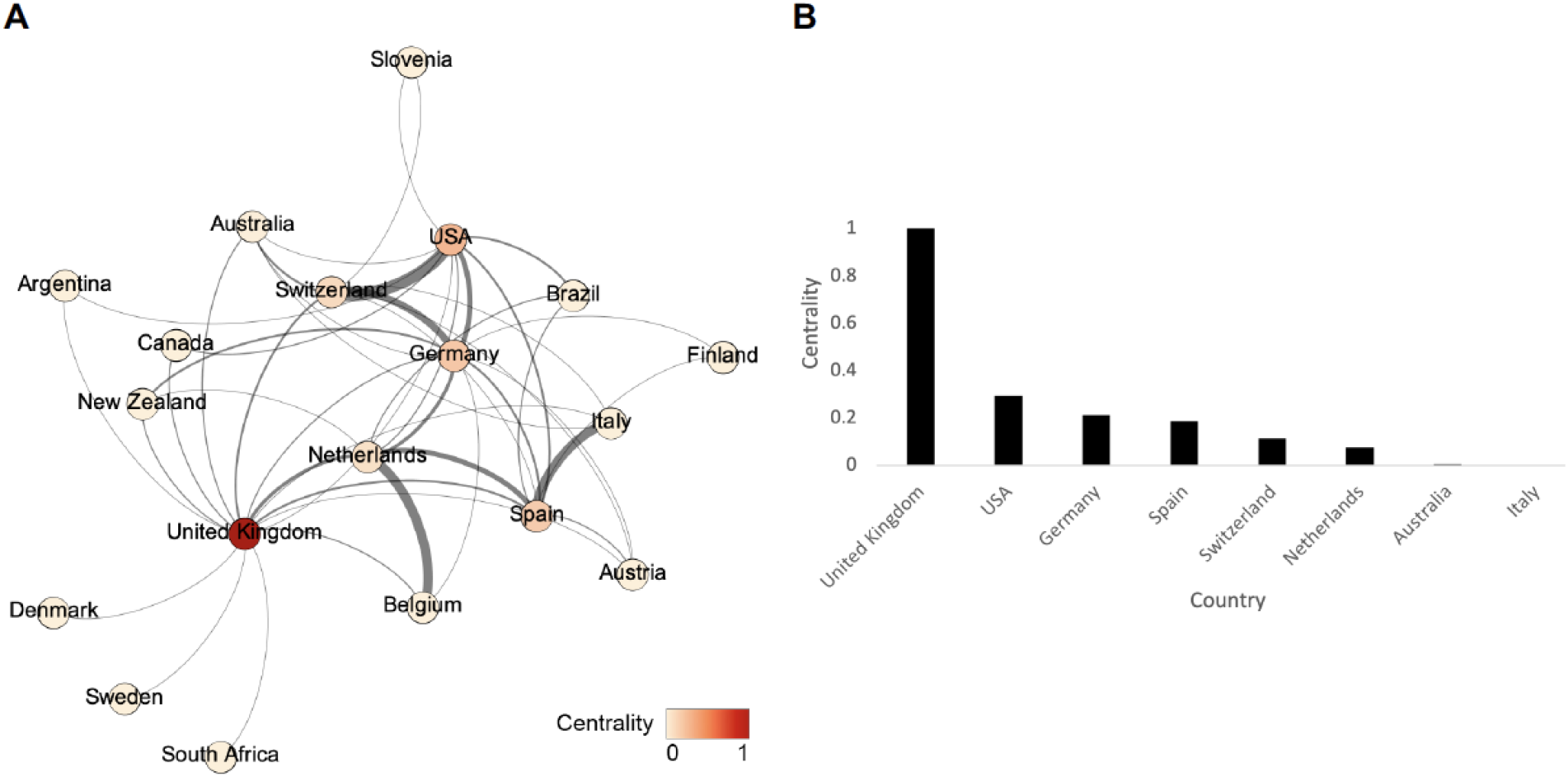
Country collaboration network. (A) Nodes represent countries (color denotes betweenness centrality). Edges represent co-occurrence of countries in the author affiliation list (edge thickness is proportionate co-occurrence frequency). (B) Bar chart representing betweenness centrality per country.

## DISCUSSION

This appears to be only the second peer-reviewed bibliometric investigation of psychedelic clinical studies. While the first such analysis (Lawrence et al. 2021), focused on the most cited articles reporting results of clinical and pre-clinical studies of classic psychedelics, this study broadens our understanding of psychedelic research in humans by its evaluation of all clinical studies of psychedelics published from 1965-2018, including those involving the non-classic psychedelics ibogaine and MDMA. Our results indicate that after a nearly three-decade lull between the 1970s and 1990s, publications on clinical studies of psychedelics have resurged over the last two decades and continue on an upward trajectory. Most psychedelic clinical studies thus far have focused on MDMA, psilocybin, and LSD. Considerably fewer articles have been published on clinical studies of ayahuasca, DMT, and ibogaine. Of note, studies on DMT and its derivatives significantly increased over the time period of 2010-2018, representing approximately 88% of all clinical studies conducted using this substance. Watchful waiting of this trend may find increased interest in investigating DMT’s utility as a substance for treating psychiatric disorders.

A review of the top 10 most cited papers before and after 2010, demonstrates how research on psychedelics has changed drastically over the last decade. Among the 10 most cited papers from the older cohort of studies, MDMA is the subject of seven papers, which were published from 1994-2001. During this period, recreational use of MDMA in the nightclub scene grew significantly and a number of deaths involving MDMA, often due to interactions with medications such as ritonavir, gained extensive media coverage, which peaked from 2000-2002 (Ahrens 2013). Media coverage during this time often centered on the threat that the drug posed to Middle America, with a focus on the risk that its use might spread from urban centers to the suburbs and rural America. There was also considerable media coverage and research into whether MDMA caused neurotoxicity and other serious side effects, which is reflected in the top ten cited papers of the older cohort. Two of these papers (McCann et al. 1998; 1994) were investigations into MDMA’s potential for serotonergic neurotoxicity, while the remainder reported investigations of MDMA’s physical and psychological effects (Franz X Vollenweider et al. 1998; Morgan 1998; Mas et al. 1999), including MDMA’s pharmacokinetic properties (de la Torre et al. 2000) and gender differences in its subjective effects (Liechti, Gamma, and Vollenweider 2001). Only two papers from this cohort were related to therapeutic applications of psychedelics, with one being a dose-response study of dimethyltryptamine published by Strassman and colleagues (Strassman et al. 1994) and the other being “Psilocybin can occasion mystical-type experiences having substantial and sustained personal meaning and spiritual significance”(R R Griffiths et al. 2006), which is said by many to have initiated the “psychedelic renaissance.” The final most cited paper from the older cohort also dealt with psilocybin and demonstrated that its effects in humans were due to 5-HT2 (serotonin) receptor activation rather than stimulation of the dopaminergic system (F X Vollenweider et al. 1998).

The top most cited papers of the recent cohort demonstrate two fundamental shifts in psychedelic human subjects research. The first of these is a transition from research focused on potential risks of psychedelics (MDMA in particular) towards research into their therapeutic potential, with nine of the papers focused on therapeutic applications of LSD, MDMA and psilocybin (Grob et al. 2011; Roland R Griffiths et al. 2016; R. L. Carhart-Harris et al. 2016; Ross et al. 2016; Mithoefer et al. 2011; Bogenschutz et al. 2015; Roland R Griffiths et al. 2011; Johnson et al. 2014; Gasser et al. 2014). The other striking change is the ascendance of psilocybin in psychedelic research, with that compound being the focus of eight of the most cited articles. These articles are wide ranging, including a functional imaging study of psilocybin’s psychological effects (R. L. Carhart-Harris et al. 2012), as well as studies of psilocybin-assisted therapy’s effects on psychologically healthy individuals (Roland R Griffiths et al. 2011), patients with depression (R. L. Carhart-Harris et al. 2016), patients experiencing psychological distress due to cancer (Grob et al. 2011; Roland R Griffiths et al. 2016; Ross et al. 2016), patients with alcohol use disorder (Bogenschutz et al. 2015), and patients with tobacco use disorder (Johnson et al. 2014). Rounding out the recent cohort are a study of LSD-assisted psychotherapy’s effects on anxiety associated with life-threatening disease (Gasser et al. 2014) and MDMA-assisted psychotherapy’s effects on treatment-resistant posttraumatic stress disorder (Mithoefer et al. 2011).

As of 2018, articles reporting on psychedelic clinical studies were primarily published in journals with a biological or pharmacologic focus. However, with recent results of clinical trials of psilocybin-assisted therapy for depression (R. Carhart-Harris et al. 2021) and MDMA-assisted therapy for PTSD (Mitchell et al. 2021) published in high profile general medical journals such as the *New England Journal of Medicine* and *Nature Medicine*, future bibliometric analyses may find that the types of journals publishing articles of psychedelics has changed.

Notably, our findings demonstrate that investigators at a small number of academic research institutions in the United States, Switzerland, Spain and a few other European countries are conducting the vast majority of psychedelic clinical research at this time. Two of these, Imperial College in the United Kingdom (O’Hare 2019) and Johns Hopkins University (Lewis 2020) in the United States, have opened research centers dedicated to the study of psychedelics in the last two years and appear poised to engage more deeply in this work. Additional centers in the United States have also recently opened at other academic institutions including Mount Sinai Health System (Steffen 2021), New York University (NYU Langone Health 2021), and the University of California at Berkeley (Anwar 2020), suggesting growing academic interest in working with psychedelics despite a lack of U.S. National Institutes of Health (NIH) funding for psychedelic-assisted therapy clinical trials (Barnett, Parker, and Weleff 2021). Scientific funding bodies in the other countries such as Australia, Canada, Israel, New Zealand and the United Kingdom have been more supportive of this research. However, NIH has recently joined them by providing funding for a study of psilocybin-assisted therapy for tobacco use disorder (Johns Hopkins Medicine Newsroom 2021).

The network analyses applied to inter-country collaborations suggests that psychedelic researchers in the United Kingdom have more diverse collaborations by working with collaborators in a large number of countries. Further, the country occupies a unique position in the network by connecting nodes of researchers who would otherwise be disconnected (Denmark, Sweden, and South Africa). The graph also reveals several strong collaborations between certain countries, such as Switzerland-USA-Germany, Belgium-Netherlands, and Italy-Spain. Intriguingly, the countries that account for the highest research output (data available in supplement) were not necessarily the most collaborative internationally (Figure 3B).

With philanthropists (Psychedelic Science Funders Collaborative 2017) and the biotechnology industry (Taylor 2021) expressing growing interest in developing psychedelic treatments and more governments funding this research, we may see the entry of a large number of new investigators and institutions into medicinal psychedelic research in coming years. Hopefully, this will produce an infusion of unique perspectives into this complex area of study, catalyzing the scientific community’s efforts to better understand these fascinating compounds and harness them as treatments.

### Limitations

The primary limitation of this study is the fact that only articles published in journals indexed in PubMed and WoS were captured in our search. Relevant older journal articles may not have been indexed, likely skewing our data towards more recent publications. Additionally, our study’s scope was limited to articles addressing clinical studies of psychedelics. Thus, articles describing basic science investigations into psychedelics were excluded, meaning that we could not examine the contributions of researchers helping the scientific community understand psychedelics from this crucial perspective.

## Data Availability

Data used in this analysis has been uploaded and made publicly available on the following repository: https://doi.org/10.6084/m9.figshare.17054405.v1
Other information can be obtained by contacting Dr. Weleff.

## Data availability statement

Data used in this analysis has been uploaded and made publicly available on the following repository: https://doi.org/10.6084/m9.figshare.17054405.v1 Other information can be obtained by contacting Dr. Weleff.

