## Supplementary Information for "Bibliometric Analysis of Academic Journal Articles Reporting Results of Psychedelic Clinical Studies"

**Table S1.** Descriptive statistics, including information on article citations, authors and author collaborations.

| Search timespan | 1965-2018 |
|---:|---:|
| Journals | 106 |
| Articles | 320 |
| Mean ± SD years since publication | 17.2 ± 14.5 |
| Median years since publication | 12 |
| Mean ± SD articles published per year | 5.9 ± 6.4 |
| Mean ± SD citations per article | 61.5 ± 72.2 |
| Mean citations per year per article | 5.2 |
| Authors | 900 |
| Authors per article | 2.8 |
| Author appearances in total | 1899 |
| Authors of single-authored articles | 17 |

**Figure S1.** Number of psychedelic clinical study articles published by journal from 1965-2018

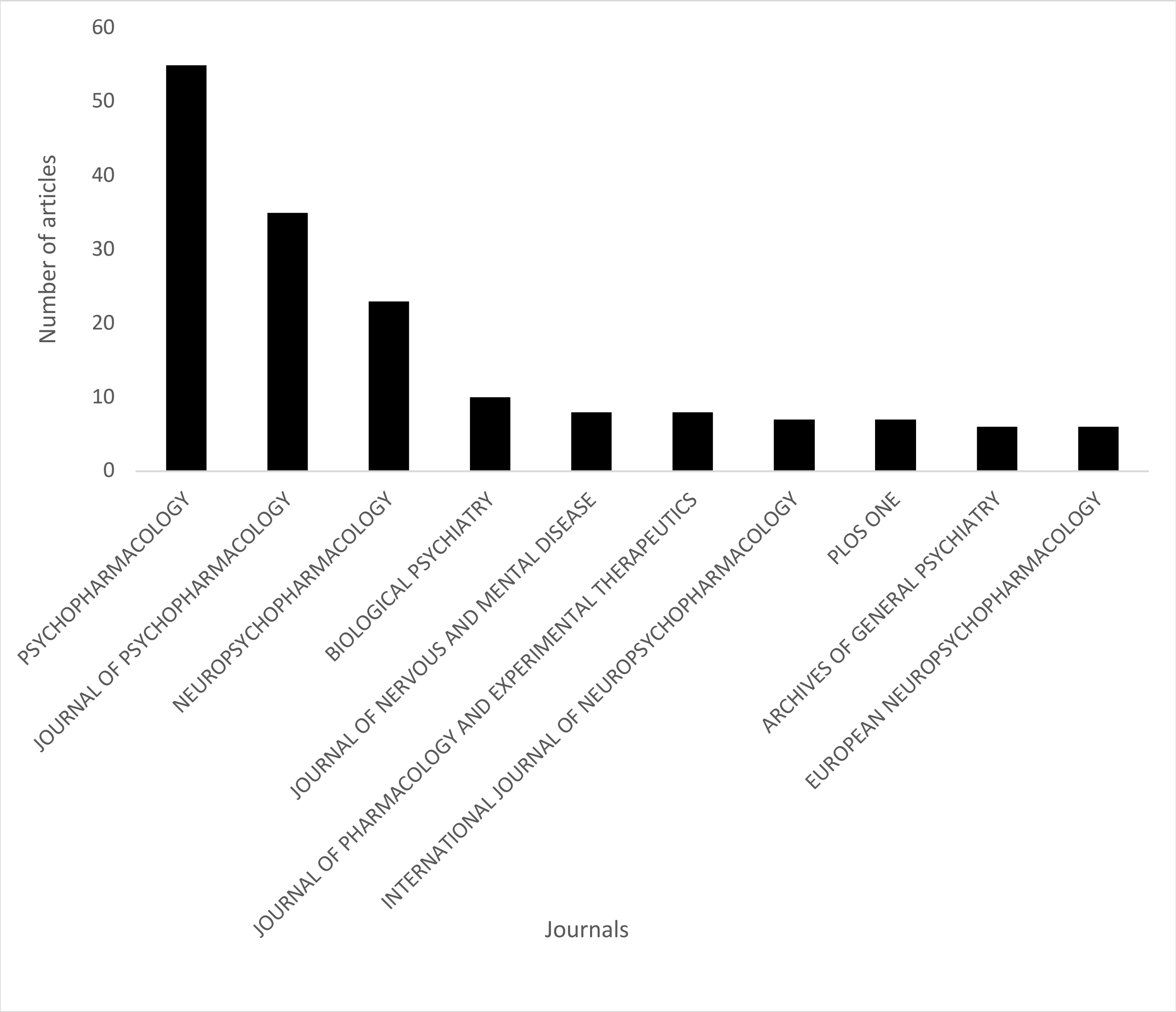

**Figure S2.** Total citations of psychedelic clinical study articles published from 1965-2018 categorized by journal of publication

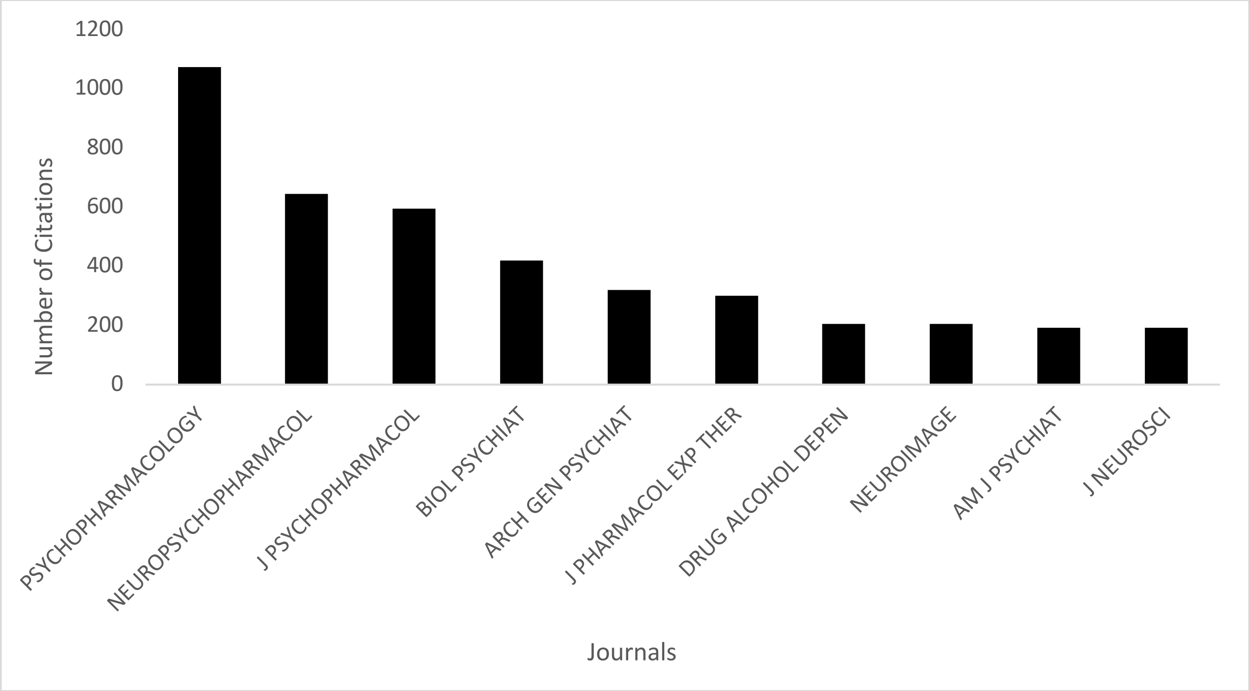


**Figure S3.** Academic institutions of corresponding authors of psychedelic clinical study articles published from 1965-2018

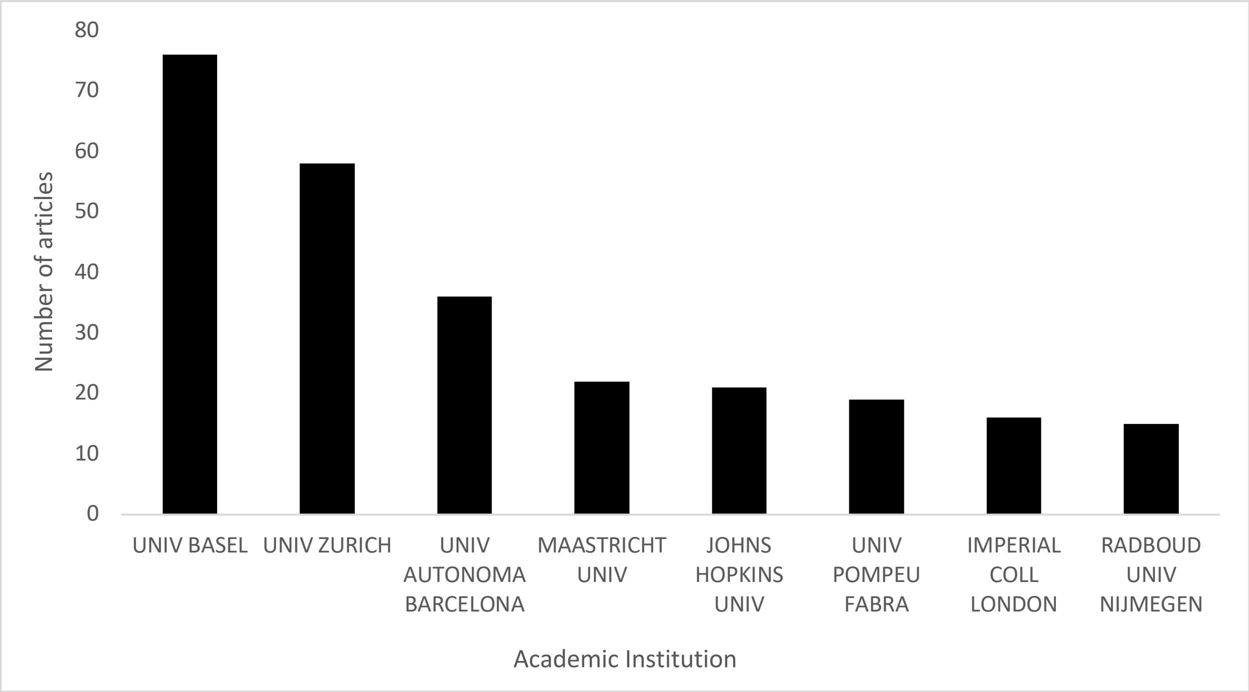

**Figure S4.** Countries of corresponding authors of psychedelic clinical study articles published from 1965-2018

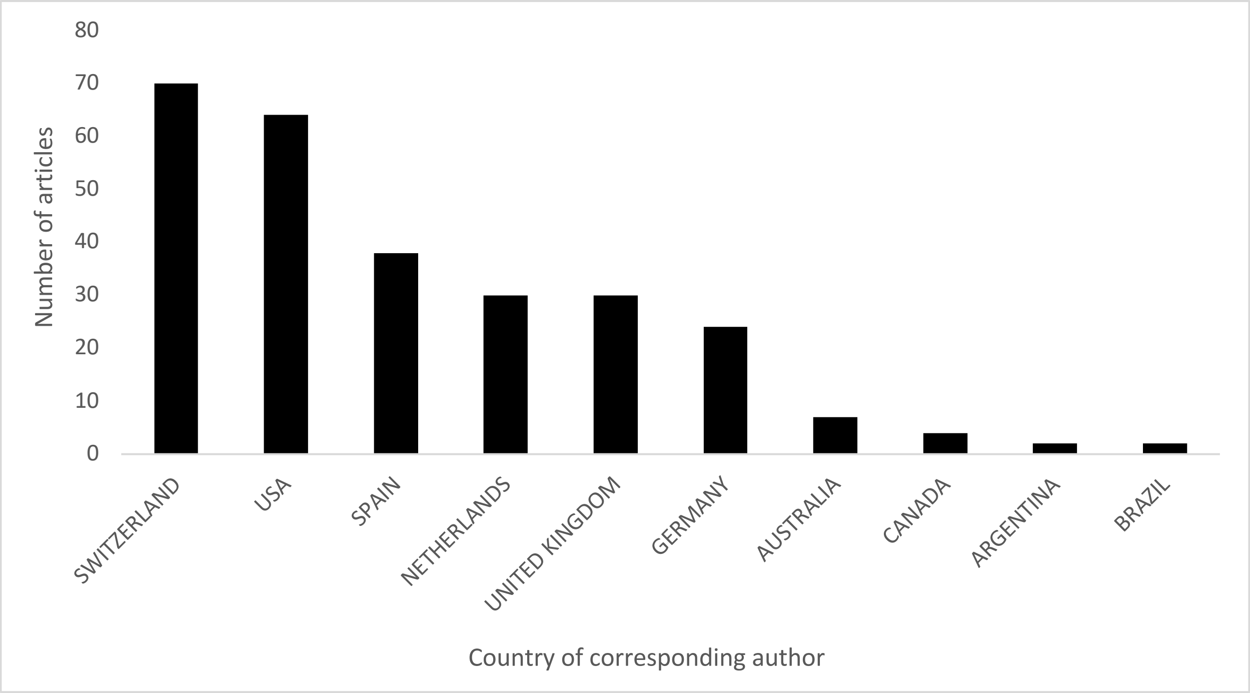


**Figure S5.** Corresponding author countries with the most citations of psychedelic clinical study articles published from 1965-2018

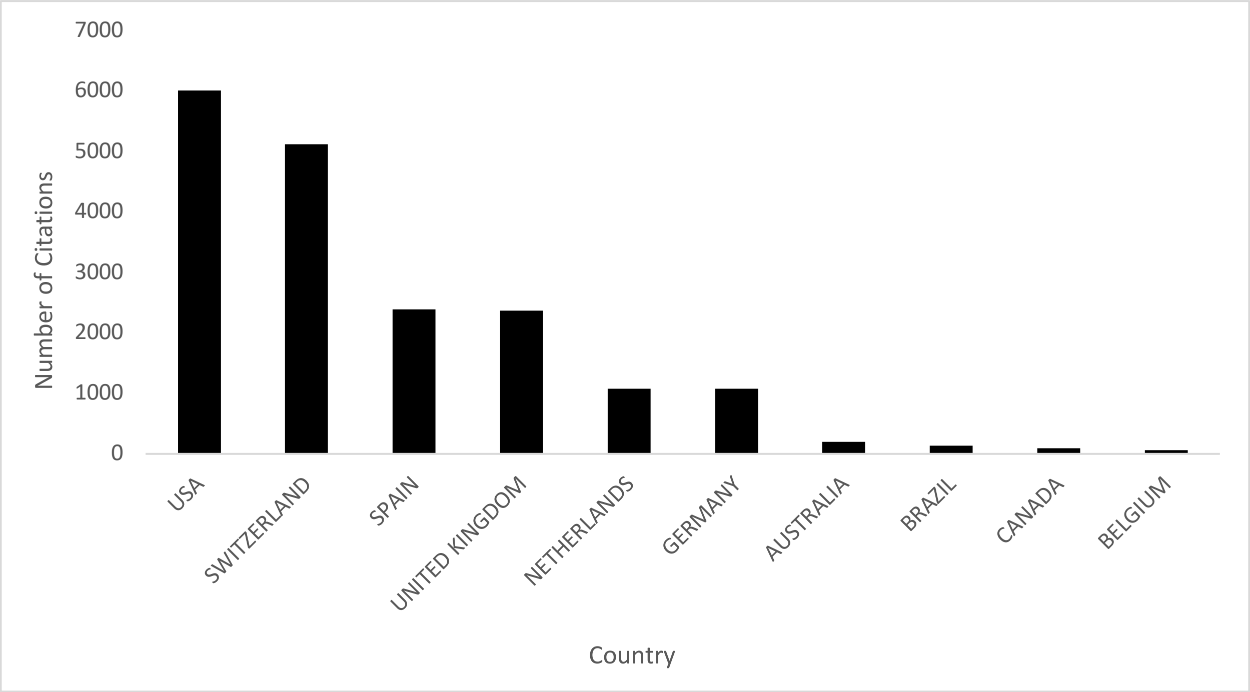
